# Agentic Generative Artificial Intelligence System for Classification of Pathology-Confirmed Primary Progressive Aphasia Variants

**DOI:** 10.1101/2025.10.28.25338977

**Authors:** Chiara Gallingani, Zachary A Miller, Maria Luisa Mandelli, Howard J Rosen, Zoe Ezzes, Mia Lin, Diana Rodriguez, Lea T Grinberg, Salvatore Spina, William W Seeley, Bruce Miller, Maria Luisa Gorno-Tempini, Pedro Pinheiro-Chagas

## Abstract

**Importance:** Accurate clinical and pathological diagnoses are essential in neurodegenerative diseases, especially given the emergence of pathology-specific disease-modifying therapies. However, diagnostic accuracy remains challenging due to heterogeneous clinical presentations, complexity of integrating multimodal data, and limited access to multidisciplinary expertise. Primary Progressive Aphasia (PPA) exemplifies these challenges, requiring specialized clinical, neuropsychological, and imaging evaluations. Generative artificial intelligence (AI), powered by large language models, may offer scalable diagnostic support in this context.

**Objective:** To evaluate the diagnostic performance of an agentic generative AI system in classifying prototypical PPA cases by clinical syndrome and underlying pathology.

**Design:** Retrospective diagnostic validation study using a multi-agent generative AI architecture simulating expert-level reasoning.

**Setting:** Single tertiary academic referral center (University of California San Francisco, Memory and Aging Center).

**Participants:** Fifty-four individuals with a definite diagnosis of PPA and post-mortem confirmation (18 semantic [svPPA], 17 logopenic [lvPPA], 19 nonfluent [nfvPPA]), selected as prototypical cases with congruent clinical, imaging, and pathological profiles.

**Exposure:** Multimodal input data, including clinical notes, neuropsychological and language assessments, and MRI brain images, were processed through a multi-agent architecture. The system generated diagnostic predictions under two conditions: (1) open-ended diagnosis from a set of 15 neurodegenerative clinical syndromes; (2) constrained classification of PPA variant and underlying neuropathology.

**Main Outcomes and Measures:** Generative AI system diagnostic accuracy for clinical syndrome and pathology, based on expert clinical diagnoses and post-mortem confirmations as gold standard.

**Results:** In the open-ended setting, the system correctly identified PPA in 49 of 54 cases (90.7%, chance level=6.7%). When constrained to PPA, it achieved 100% accuracy for svPPA and nfvPPA, and 94.1% for lvPPA as primary prediction. Neuropathological predictions were most accurate for FTLD-TDP type C (100%) and FTLD-4R tau (100%), and high for Alzheimer’s disease (94.4%). The full diagnostic pipeline of all 54 cases was completed in under 10 minutes.

**Conclusions and Relevance:** The AI system demonstrated expert-level performance in classifying prototypical PPA cases, integrating multimodal data and mirroring specialist reasoning. Its speed and accuracy support its potential role in extending access to specialized diagnostic expertise, particularly in non-tertiary settings. Further validation in larger and more heterogeneous populations is warranted.

## Introduction

Early diagnosis and accurate pathological prediction represent critical needs in the field of neurodegenerative dementias. Timely identification and multidisciplinary assessment have been shown to improve quality of life and reduce long-term healthcare costs^1^, while anticipating the underlying pathology is essential for ensuring access to emerging disease-modifying therapies. However, more than 50% of dementia diagnoses are delayed until moderate or advanced stages, with even greater delays among minorities^2^. Achieving diagnostic accuracy is challenging – even in tertiary centers – due to heterogeneous clinical presentations, need for extensive data collection and integration, and rapidly evolving medical knowledge.

Additionally, the population is aging worldwide, and the number of US dementia cases are expected to double by 2060, reaching 1 million annually^3^. When approaching neurodegenerative diseases, primary care providers often need to refer patients to specialists^4^. Consequently, the gap between the number of individuals requiring specialized care and the availability of trained professionals and healthcare resources is increasing^5,6^, as well as wait times for reaching specialists especially in rural areas compared to urban regions^7^.

In this context, generative artificial intelligence (AI) powered by large language models (LLMs) may represent a useful and scalable tool. AI systems using LLMs are a rapidly evolving class of AI trained on vast databases of text, with great potential in understanding and generating natural language. In the clinical setting, LLMs can process and synthesize large amounts of multimodal data – including clinical texts and reports, audio recordings, images – providing a unified framework^8–10^. Moreover, LLMs have shown promise in clinical reasoning, reaching similar performance of general physicians^11^, and retrieval-augmented generation has been used to ground LLMs to the current diagnostic criteria^12^. Advances in LLM-based AI systems hold potential to improve diagnostic accuracy and expand access to expert-level care, by supporting the clinical workflows through the application of updated medical knowledge^13^.

The evaluation and validation of these systems on real word data and authentic clinical scenario are needed. One opportunity is represented by Primary Progressive Aphasia (PPA)^14^, a group of neurodegenerative syndromes characterized by predominant language impairment, which constitutes a valuable model for both diagnostic challenge and need for multidisciplinary expertise.

Three main variants of PPA are well recognized in English-speaking populations, each with relatively distinct clinico-anatomical and pathological profiles in prototypical presentations^14^. The semantic variant (svPPA) is defined by impaired confrontation naming and single-word comprehension, typically associated with asymmetric anterior temporal lobe atrophy and frontotemporal lobar degeneration (FTLD) with TDP-43 type C pathology^15,16^. The nonfluent variant (nfvPPA) presents with effortful speech, agrammatism and/or motor speech deficits, linked to atrophy in the inferior frontal gyrus, premotor cortex, and supplementary motor area, and to FTLD 4R-tauopathies (corticobasal degeneration [CBD] and progressive supranuclear palsy [PSP])^15,17^. The logopenic variant (lvPPA), typically associated with Alzheimer’s disease (AD) pathology, involves phonological processing difficulties, word-finding pauses, and parietotemporal atrophy^15,18^.

Despite these well-defined criteria, accurate and early subtyping of PPA variants remains challenging. In primary neurology settings, it is often inaccessible, due to limited multidisciplinary resources, such as speech and language pathologists, and lack of time and confidence with complex multimodal information. In specialized clinics, the diagnosis still requires a huge effort in terms of time, staff, and continuous medical education.

The aim of this study was to evaluate the performance of an agentic generative AI system, developed at the Memory and Aging Center (MAC) of the University of California San Francisco (UCSF). It is a decision support tool designed to synthesize multimodal clinical data and incorporate MAC’s expertise to aid diagnostic prediction and clinical workflows^19^. We tested the feasibility and utility of its implementation in classifying a set of well-characterized, typical PPA cases with available clinical, MRI, and neuropathological data. We hypothesize that the current available AI technology implemented in a specifically designed multi-agentic system can accurately classify each case into the correct syndromic and pathological category, aligning with expert-level clinical reasoning.

## Methods

### 2.1 Case selection

The UCSF MAC database was screened for PPA cases, fulfilling a definite diagnosis of svPPA, nfvPPA, or lvPPA^14^. Each case had undergone a comprehensive neurological evaluation, documented in detail within a clinical note, as well as extensive neuropsychological, language, and speech assessments. Diagnoses were performed for each case through the consensus of an expert team. Specifically, the consensus is reached during a one-hour multidisciplinary conference where data collected are reviewed and discussed. Eligible participants had available MRI scan demonstrating atrophy patterns consistent with their clinical syndromes and neuropathological post-mortem confirmation (i.e., svPPA: FTLD-TDP-C, lvPPA: AD, nfvPPA: FTLD-4R tauopathy, either CBD or PSP). To align participant selection with the aim of a proof-of-concept design, we included only prototypical cases, with congruent imaging findings and confirmed neuropathology. All participants provided written informed consent, under UCSF Human Research Committee approval.

### 2.2 Data Modalities and Preprocessing

The AI system received multimodal input – including clinical notes, neuropsychological test results, and MRI-derived atrophy scores – which was preprocessed into tabulated data (Figure 1).

**Figure 1.**
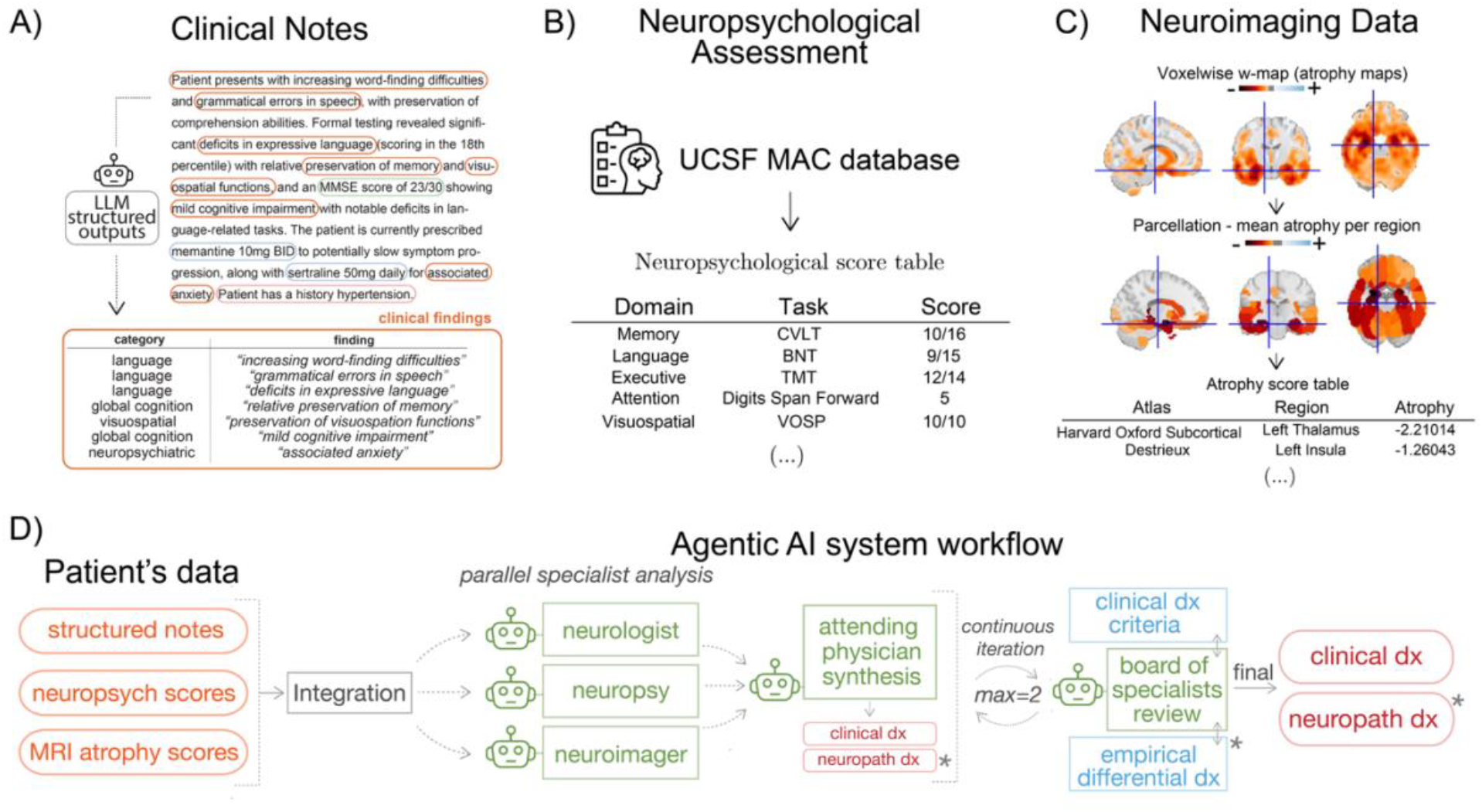
Input data preparation for the agentic generative AI system. (A) Clinical notes were first processed to extract detailed clinical information, structured into four domains: (i) clinical findings, including specific clinical symptoms and signs, predefined neurological/ neuropsychiatric categories, temporal details, severity, supporting evidence, and confidence levels, (ii) quantitative measurements, including measurement values, units, test types, test names, cognitive/behavioral domains, supporting evidence, and confidence levels, (iii) medications, including medication names, dosages, frequencies, statuses, supporting evidence, and confidence levels, and (iv) medical history, including documented conditions, onset details, status, treatment history, supporting evidence, and confidence levels. Only clinical findings were input in the model. (B) Cognitive and language scores collected from the neuropsychological and speech and language evaluations and stored in the UCSF MAC database were extracted and structured in organized tables, where every task and its corresponding score was divided into five domains. (C) Preprocessed brain MRI scans produced w-maps which were parcellated using three different atlases to obtain subject’s atrophy score tables. D) Agentic AI System Workflow. The system passes the integrated data to three specialized agents (the behavioral neurologist, the neuropsychologist, and the neuroimaging agent). An attending physician LLM combines their analyses to predict diagnoses, which are then reviewed by an LLM board of specialists for final approval. The board has access and can consult if needed to the MAC diagnostic criteria and the empirical differential diagnosis. * Only available for the Constrained PPA Variant Classification.

#### 2.2.1 Clinical Notes

Clinical data was extracted from clinical notes using a structured information extraction pipeline powered by OpenAI’s gpt-4.1 (version 2025-04-14), accessed via the UCSF Versa platform, which is HIPAA-compliant and approved for use with P4 identifiable health data (see Supplementary Material). The extraction process employed predefined JSON output schemas through Pydantic validation to ensure consistency and completeness. Information was organized into four domains: (1) *clinical findings*, including anamnestic history, symptoms, and signs with temporal details and severity judgements, (2) *quantitative measurements*, encompassing cognitive test scores, (3) *current medications*, with dosages and frequencies, and (4) *medical history,* with past conditions and treatment records. To prevent diagnostic bias, we employed two strategies: 1) any explicit diagnostic impressions or conclusions in the original note were removed and 2) we only provided the AI system with *clinical findings*, since the other fields could be revealing of the diagnosis (e.g. anticholinesterase inhibitors for AD).

#### 2.2.2 Neuropsychological Assessment

The neuropsychological evaluation comprised a standardized battery of cognitive assessments^20^ along with a speech and language battery designed specifically for PPA evaluation^15^. The language assessment tested speech production, comprehension, confrontation naming, repetition, and reading. We only included the speech and language battery in the *Constrained PPA Variant Classification* (see below 2.5), to minimize bias towards an open-ended diagnosis of PPA. Raw test scores were converted to standardized formats and organized into domain-specific categories encompassing memory (episodic, semantic, working memory), language (fluency, naming, comprehension, grammar), executive function (planning, inhibition, cognitive flexibility), attention and processing speed, and visuospatial abilities.

#### 2.2.3 Neuroimaging Data

Structural brain MRI scans underwent preprocessing of T1 sequences to generate patient-specific whole-brain atrophy maps (w-maps), as previously described^21,22^ and detailed in Supplementary Materials. W-maps represents voxel-wise z-scores relative to a group of healthy controls after adjusting for age, sex, and total intracranial volume. A z-score below –1.65 represents significant atrophy relative to controls. The neuroimaging pipeline also implemented a multi-atlas parcellation approach using the Destrieux cortical atlas (150 regions), the Automated Anatomical Labeling (AAL) atlas (116 regions), and the Harvard-Oxford Subcortical atlas (21 regions). For each atlas, ‘NiftiLabelsMasker’ from the Nilearn library was employed to extract the mean z-score within each parcel. This approach provided comprehensive coverage of cortico-subcortical structures, enabling detailed analysis of region-specific atrophy patterns of PPA variants, including hemis-pheric asymmetries and structure-function relationships.

### 2.3 Agentic Framework Architecture: Multi-Agent Clinical Assessment System

The AI system implemented a multi-agent architecture simulating the real multidisciplinary clinico-pathological diagnostic workflow used at UCSF MAC, in which specialized AI agents represented different medical expertise domains (Figure 1, see also Supplementary Materials for additional technical implementation). All agents were programmed through detailed prompt engineering reflecting clinical logic and decision-making, utilizing OpenAI’s o4-mini model (version 2025-04-16) with high reasoning effort, to replicate specialist-level reasoning throughout the agentic workflow.

#### 2.3.1 Behavioral neurologist, neuropsychologist, and neuroimaging agents

The workflow commenced with parallel assessment by three specialist agents – modeled after a behavioral neurologist, a neuropsychologist, and a neuroimaging specialist –, each receiving identical multimodal patient data but analyzing it through their respective domain-specific expertise and prompting strategies. The *behavioral neurologist agent* focused on clinical syndrome recognition, analyzing symptom patterns, disease progression, and particularly language and communication features. The *neuropsychologist agent* interpreted neuropsychological test data, identifying specific patterns across language, memory, executive function, attention, and visuospatial domains. The *neuroimaging agent* examined structural MRI data, assessing atrophy patterns and functional implications. Each agent generated detailed, domain-informed assessments without producing explicit diagnostic labels.

#### 2.3.2 Attending Physician Agent

Following parallel specialist assessments, an integrator agent synthesized the three analyses into a comprehensive diagnostic formulation. This agent was designed as a “*senior neurologist who specializes in identifying the most diagnostically decisive clinical features from multidisciplinary assessments.*” The prompt emphasized focus on 3-5 key features that most strongly distinguished each presentation, selecting findings that: (1) converged across domains, (2) represented distinctive or unusual presentation features, (3) possessed discriminative value for differential diagnosis, and (4) avoided common or overlapping features in favor of case-specific distinguishing characteristics. This agent provided structured outputs including a *primary* and *differential* clinical syndrome diagnosis and a *primary* and *differential* pathology prediction.

#### 2.3.3 Expert Board Review Agent

The diagnostic formulation underwent review by an expert board agent simulating a multidisciplinary review panel. This agent was provided with diagnostic criteria and empirical differential diagnosis database (only for the Constrained PPA Variant Classification, see Experimental design and Supplementary Materials) and evaluated diagnostic formulations for accuracy, consistency, and adherence to established criteria. The board agent systematically evaluated: (1) synthesis quality and convergent findings among specialists, (2) diagnosis alignment with established criteria, (3) differential diagnosis considerations, (4) neuropathological prediction accuracy based on clinicopathological correlations, and (5) overall concordance.

#### 2.3.4 Iterative Refinement Process

The system incorporated an iterative refinement mechanism allowing up to two revision cycles. When the board status indicated “Needs Revision,” a refinement agent re-analyzed the case incorporating specific feedback from the board review. This refinement agent was tasked with directly addressing board concerns while maintaining comprehensive diagnostic assessment and ensuring anatomical-clinical concordance according to established criteria.

### 2.5 Two-Stage Experimental Design

#### 2.5.1 Stage 1: Open-Ended Neurodegenerative Diagnosis

In the initial experimental condition, the AI system performed open-ended diagnosis without prior knowledge of PPA, selecting from a set of 15 neurodegenerative conditions including AD, behavioral variant Frontotemporal Dementia (bvFTD), Corticobasal Syndrome, PSP, Dementia with Lewy Bodies, and PPA among others. This condition tested the system’s ability to identify PPA within numerous neurodegenerative diseases, simulating real-world clinical scenarios where the specific syndrome is unknown.

#### 2.5.2 Stage 2: Constrained PPA Variant Classification

In the constrained experimental condition, the system was informed on PPA diagnosis and tasked with variant classification and neuropathological prediction. The system was also provided with the specific language battery used for assessing PPA, in addition to the general neuropsychological battery. The system classified cases into svPPA, lvPPA, or nfvPPA and predicted underlying pathology from four categories: AD pathology, FTLD-TDP-C, FTLD-4R-TAU PSP, and FTLD-4R-TAU CBD. This condition evaluated the system’s diagnostic precision when focused specifically on PPA subtyping and pathological prediction.

## Results

### 3.1 Sample characteristics

A total of 54 PPA cases were included: 18 svPPA (all with TDP-C pathology), 17 lvPPA (all with AD pathology), and nfvPPA cases (10 with CBD, 9 with PSP pathology). Demographics and global cognitive measures for the overall sample and each diagnostic group are reported in Table 1. Subjects with nfvPPA were significantly older than those with lvPPA; subjects svPPA had the longest disease duration and poorer scores on general cognition measures compared to nfvPPA.

**Table 1:** Participant demographics and general cognition.

|  | **Total sample**<br>**n = 54** | **lvPPA**<br>**n = 17** | **nvPPA**<br>**n = 19** | **svPPA**<br>**n = 18** | **T stat** | **p-value** |
| --- | --- | --- | --- | --- | --- | --- |
| **Age (years)** | 65 (58.8-71.8) | 59 (57-68) | 72 (65-75) | 64.5 (58.5-68) | 11.09 | **0.004<sup>a</sup>** |
| **Gender (male)** | 22/54 (40.7%) | 4/17 (23.5%) | 10/19 (52.6%) | 8/18 (44.4%) | 3.30 | 0.192 |
| **Handedness (right)** | 43/54 (79.6%) | 11/17 (64.7%) | 19/19 (100%) | 13/18 (72.2%) | 7.80 | 0.020<sup>d</sup> |
| **Ethnicity (White, Latino/Hispanic, Japanese)** | 52,1,1 | 17,0,0 | 18,0,1 | 17,1,0 | 3.88 | 0.423 |
| **Education (years)** | 16 (15.2-18) | 17 (16-18) | 16 (13-18) | 16 (16-17.8) | 4.13 | 0.127 |
| **Disease duration (years)** | 4 (2.25-6) | 4 (2-5) | 3 (2-4.5) | 6 (4-8) | 10.58 | **0.005<sup>b,c</sup>** |
| **MMSE (30)** | 25 (19-27.2) | 22 (19-26) | 27 (25-27.5) | 24 (17-28.2) | 4.64 | 0.098 |
| **CDR total score (3)** | 0.5 (0.5-1) | 0.5 (0.5-0.63) | 0.5 (0.5-0.5) | 1 (0.5-1) | 6.77 | **0.034<sup>b</sup>** |
| **CDR box score (18)** | 3.5 (1.5-4.5) | 4 (2.25-4.12) | 2 (1-2.25) | 4 (3.5-5.88) | 9.64 | **0.008<sup>b</sup>** |

Descriptives are reported as median (IQR) for continuous variables, or frequency (%) for categorical variables. Group comparisons were performed using Kruskal–Wallis, or chi-square tests, followed by Bonferroni-corrected post hoc tests where appropriate. a = nvPPA vs lvPPA, b = nvPPA vs svPPA, c = lvPPA vs svPPA, d = no significant post hoc comparisons.

### 3.2 Diagnostic Performance of AI system

#### 3.2.1 Stage 1: Open-Ended Neurodegenerative Diagnosis

In the initial experimental condition of an open-ended diagnosis among 15 neurodegenerative conditions (Figure 3, panel A-B), the AI system correctly classified 49 of 54 cases (90.7%) as PPA, with five misdiagnosed cases: 2 AD, 2 bvFTD, 1 PSP. When considering both the primary and differential matches, the AI system correctly identified 52 of 54 cases (96.3%). The expected chance level, given the 15 diagnostic categories, is approximately 6.7%.

**Figure 3.**
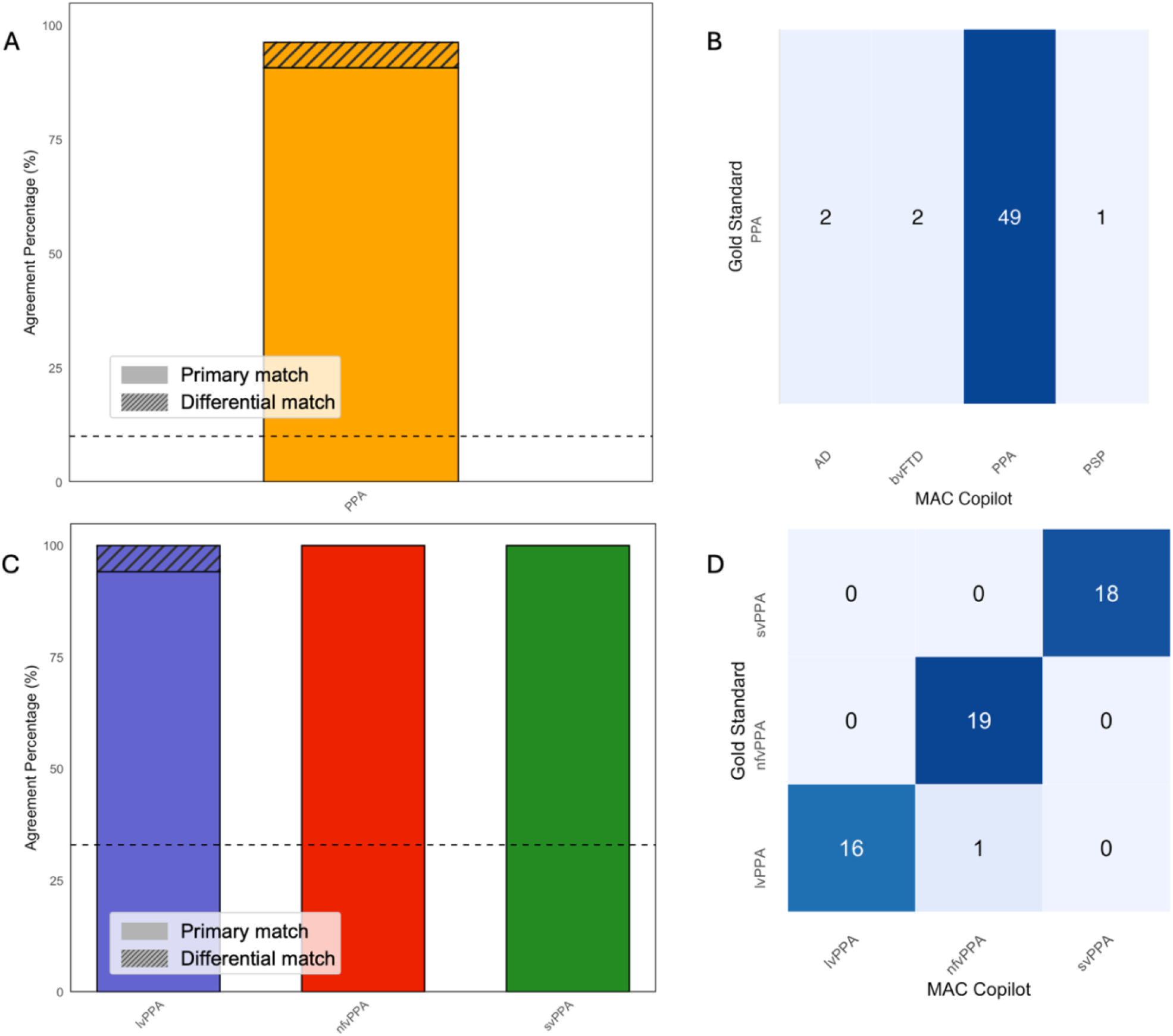
AI system performance in predicting clinical diagnosis. (A) General accuracy in identify PPA among 15 possible diagnoses. (B) Distribution of open-ended diagnoses, showing 5 misdiagnoses among 54 patients (2 AD, 2 bvFTD, 1 PSP). (B) Results in the constrained setting, with 100% accuracy for svPPA and nfvPPA, and 94.1% for lvPPA as primary diagnosis (100% considering also the differential match). (C) Confusion matrix confirming strong accuracy for nfvPPA (19/19 correct) and svPPA (18/18 correct), with 1 misdiagnosis among 18 lvPPA (1 nfvPPA). Dashed lines in the histograms represent the expected chance level of choice.

#### 3.2.2 Stage 2: Constrained PPA Variant Classification

When explicitly informed about the PPA diagnosis and asked to specify the variant (Figure 3, panel C-D), all previously misdiagnosed participants were correctly reassigned to the PPA category. Overall, all svPPA (19/19) and nfvPPA (19/19) cases were correctly classified. Among lvPPA cases, 16/17 (94.1%) were correctly classified as the primary match, while 17/17 (100%) were correctly listed when considering also the differential match. The single misclassified lvPPA subject was labeled as nfvPPA.

In this second stage, the model was also prompted to produce a pathological prediction (primary match and differential match), based on clinico-pathological correlations (Figure 4). In this setting, the AI system achieved perfect accuracy for FTLD-TDP-C pathology (100%), FTLD-4R-TAU category (100%), and AD pathology (100%) already with the primary match. When the given possible neuropathological predictions specifically distinguished between CBD and PSP within 4R-tauopathy, the accuracy was still high for CBD cases (80.0% primary match), but only moderate in PSP cases (66.7%). For both conditions, it reached the 100% accuracy when considering the differential match. Human-based predictions of underlying pathology, which were retrieved when available from clinical notes, yielded an accuracy of 88.2% for AD, 80% for FTLD-TDP, 42.9% for PSP, and 60% for CBD (see Supplementary table 1 for details).

**Figure 4.**
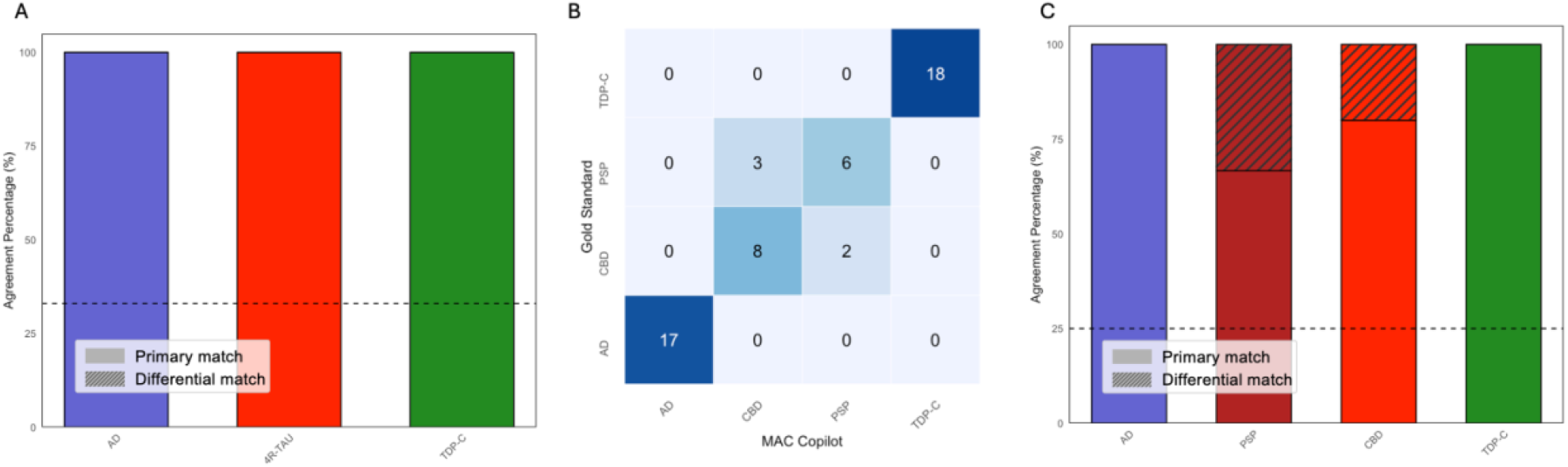
AI system performance in predicting underlying neuropathology. (A) Performance with grouped 4R-tauopathies (CBD and PSP) reached 100% accuracy for each category. (B) Confusion matrix for detailed neuropathological diagnosis (distinguishing between CBD and PSP) shows perfect accuracy for AD and TDP-C (18/18), with 3/9 PSP and 2/10 CBD misclassified as the other 4R-tauopathy. (C) Plots for the detailed neuropathological diagnosis show an accuracy of 66.7% for PSP and 80.0% for CBD in the primary match, both reaching 100% when including differential matches. Dashed lines in the histograms represent the expected chance level of choice.

The full diagnostic pipeline for all 54 cases running in parallel was completed in approximately 10 minutes. To assess the consistency of predictions, we ran the complete model 10 times for each patient; the output remained stable in all but two runs, where two patients were classified differently than in the original analysis, and in both instances the predictions were incorrect.

## Discussion

We developed and evaluated accuracy of an agentic AI system in reproducing diagnosis of prototypical PPA clinico-pathological variants. We structured a proof-of-concept design using a cohort of well-characterized and typical PPA cases, for which consensus clinical diagnosis and neuropathological post-mortem confirmation were available, and tested the ability of our agentic AI system to reproduce the expert diagnostic outcome. We showed that this ad-hoc-designed generative AI system, informed with the same multimodal data available to human experts, can accurately classify both clinical variants and underlying neuropathology. The AI system not only produced accurate diagnostic predictions but also generated plausible narrative outputs. Furthermore, a careful qualitative review of the generated reports confirmed that in every case, the system effectively integrated the multimodal data to support its diagnostic and pathological predictions. Therefore, this AI system reaches similar conclusions to world-class specialist humans based on the same cues, suggesting a role in streamlining and scaling access to specialized knowledge.

A key strength of this AI system is its speed and possible scalability for multiple clinical purposes. Once the system was constructed, the entire diagnostic process for all 54 cases was completed in under 10 minutes. This timeline sharply contrasts with the time-consuming and limited availability of the expert knowledge necessary to evaluate multimodal information. The diagnostic workflow used at the UCSF MAC that inspired the architecture of the agentic AI model requires at least 4 expert healthcare professionals, including a neurologist fellow for the anamnestic interview and the neurological exam, a neuropsychologist for the cognitive assessment, a speech and language pathologist for the speech and language evaluation, and an attending neurologist who presides over the final multidisciplinary conference (1 hour per patient) where collected data are reviewed, synthesized and integrated to formulate the final diagnoses. By integrating the perspectives of these multidomain experts, the tool can quickly emulate a multidisciplinary evaluation, offering a level of refinement often unavailable in less specialized settings. This capacity has important implications: only about 20% of patients with dementia currently receive specialist-level evaluation in the first year^23^. Expanding access to expert-level tools could empower less specialized clinicians and facilitate timely referral to third-level care and treatment.

Another notable strength of this AI system lies in its ability to rapidly extract and categorize relevant information from complex narrative sources, such as clinical notes. This function may be useful in multiple ways such as in training and enhancing decision-making for both experienced and less experienced clinicians. In particular, a future development of this tool may include an interactive platform to support real-time clinical evaluations, guiding users in identifying which aspects of history or examination need to be expanded. Another future application of the model may include the extraction of common symptom patterns in a specific neurodegenerative condition from clinical interviews. This information may assist the research community in identifying new relevant clinical features to further refine syndromic categorization and diagnostic criteria. This could also support the development of standardized checklists for clinical evaluation that improve diagnostic consistency and accuracy across diverse settings.

Final pathological confirmation for neurodegenerative diseases is rarely available even in highly specialized dementia clinics. Therefore, this agentic AI system trained on dataset with pathological confirmation hold the potential to provide expert pathological prediction in pan ante-mortem situations, especially for disease such as PPA variants, in which biomarkers are still not available. Our system demonstrated high-level of accuracy in predicting the three broad neuropathology of AD, TDP-C, and 4R-TAU in prototypical PPA cases, while showing a slightly lower performance in distinguishing PSP and CBD within the 4R-tauopathy spectrum. This challenge mirrors the well-documented difficulty encountered by human specialists^24^; in our sample, human accuracy for these two pathologies was also limited (42.9% for PSP, 60% for CBD), and the AI system outperformed clinicians in identifying CBD (80.0%) and PSP (66.7%) within the nfvPPA variant. Future applications of this tool could allow clinicians to submit clinical data to online available specialized AI systems and receive estimation of underlying pathological etiology.

Interestingly, in the single clinically misclassified lvPPA case, both the AI system and the first clinical evaluation argued between lvPPA and nfvPPA as differential diagnoses, based on similar cues including effortful speech and misidentification of phonological errors as motor speech impairment. Only after a further revision of the case, based also on clinical progression, expert clinicians were unanimous with the lvPPA diagnosis.

This initial study has some limitations. First, this is a proof-of-concept, first-of-its-kind study including a relatively small number of highly selected cases, limiting generalizability to broader clinical populations, particularly those with atypical or mixed presentations. Moreover, some expected differences in demographics and general cognitive measures (i.e., older age for nfvPPA subjects and longer disease duration with higher functional impairment for svPPA subjects) may represent a bias for AI classification. Second, because all cases were drawn from a single tertiary center, input data may not reflect the diversity of real-world settings. While the AI system demonstrated strong performance when provided with complete clinical information, its accuracy with limited or incomplete data needs to be further explored.

In conclusion, our specialized AI agentic system accurately identified clinical and pathological diagnoses in prototypical PPA cases, replicating expert-level performance with greater speed. By integrating multimodal data and emulating multidisciplinary expertise, it holds promise to extend specialized diagnostic training and support into clinical practice.

## Data Availability

Academic, not-for-profit investigators can request subjects, tissue and laboratory specimens, archived and imaging data, technological tools or video clips of behaviors for professional education and research for research studies from the UCSF Memory and Aging Center, but you must have Institutional Review Board approval from the UCSF Human Research Protection Program (HRPP). For more information, see https://memory.ucsf.edu/research-trials/professional/open-science.

## Author Contributions

Chiara Gallingani, Maria Luisa Gorno-Tempini, and Pedro Pinheiro-Chagas had full access to all study data and take responsibility for the integrity of the data and the accuracy of the analyses. Maria Luisa Gorno-Tempini and Pedro Pinheiro-Chagas jointly supervised the work and are considered co–senior authors.

*Conceptualization, design, and implementation of the Agentic AI system:* Pedro Pinheiro-Chagas

*Statistical analysis and Data Visualization:* Chiara Gallingani and Pedro Pinheiro-Chagas

*MRI data analysis:* Maria Luisa Mandelli, Pedro Pinheiro-Chagas

*Neuropathological diagnosis:* William W. Seeley

*Data curation:* Zachary A. Miller, Maria Luisa Mandelli, William W. Seeley, Maria Luisa Gorno-Tempini, Chiara Gallingani, Pedro Pinheiro-Chagas

*Clinical assessments:* Zachary A. Miller, Howard J. Rosen, Zoe Ezzes, Bruce Miller, Maria Luisa Gorno-Tempini

*Manuscript drafting:* Chiara Gallingani, Maria Luisa Gorno-Tempini, Pedro Pinheiro-Chagas

*Lead manuscript writing:* Chiara Gallingani

*Critical revision of the manuscript for important intellectual content:* Bruce Miller, Maria Luisa Gorno-Tempini, Pedro Pinheiro-Chagas

*Funding acquisition:* Maria Luisa Gorno-Tempini, Bruce Miller, Pedro Pinheiro-Chagas

*Administrative, technical, or material support:* Diana Rodriguez, Mia Lin

*Supervision:* Maria Luisa Gorno-Tempini, Pedro Pinheiro-Chagas

## Conflict of Interest Disclosures

The authors have no conflict of interest to declare.

## Funding/Support

Support for the UCSF Memory and Aging Center was provided by grants NIA P50-AG023501, P30-AG062422, and P01-AG019724. Additional funding was provided by R01NS050915 (to MLGT) and the UCSF Catalyst Award (to PPC).

## Role of the Funder/Sponsor

The funders had no role in the design and conduct of the study; collection, management, analysis, and interpretation of the data; preparation, review, or approval of the manuscript; or decision to submit the manuscript for publication.

## Acknowledgments

We gratefully acknowledge Rose George and Joe Hesse for their indispensable support in developing and maintaining the technical infrastructure of the UCSF Memory and Aging Center (MAC). We also thank the UCSF Tiger Team for facilitating access and use of the UCSF Versa platform. We are particularly indebted to Katherine Possin and Andreas Rauschecker for their thoughtful discussions and critical feedback on the development and design of this project.

We thank the UCSF MAC clinical team, including physicians and research coordinators, as well as the administrative staff who coordinated the research studies. Most importantly, we extend our deepest gratitude to the patients and families who participated in this research,

## Notes

### Competing Interest Statement

The authors have declared no competing interest.

### Author Declarations

The Institutional Review Board of University of California San Francisco gave ethical approval for this work (IRB #24-40959).

### Summary of Updates

Update the co-authors list, adding Lea T Grinberg and Salvatore Spina.

