## Supplementary Material for "Agentic Generative Artificial Intelligence System for Classification of Pathology-Confirmed Primary Progressive Aphasia Variants"

### Supplementary Materials

**Supplementary Table 1**. Generative AI system syndromic and neuropathological predictions for all cases, compared to human neuropathological predictions.

| Case nr. | Confirmed diagnosis | Confirmed neuropathology | AI system clinical prediction (primary match) | AI system clinical prediction (differential match) | AI system pathological prediction (primary match) | AI system pathological prediction (differential match) | Human pathological prediction |
|---|---|---|---|---|---|---|---|
| ***1*** | lvPPA | AD | nfvPPA | lvPPA | AD | FTLD-tau CBD | Not specified |
| ***2*** | lvPPA | AD | lvPPA | svPPA | AD | FTLD-TDP-C | AD |
| ***3*** | lvPPA | AD | lvPPA | svPPA | AD | FTLD-tau CBD | AD |
| ***4*** | lvPPA | AD | lvPPA | nfvPPA | AD | FTLD-tau CBD | AD |
| ***5*** | lvPPA | AD | lvPPA | nfvPPA | AD | FTLD-tau CBD | AD |
| ***6*** | lvPPA | AD | lvPPA | svPPA | AD | FTLD-TDP-C | AD |
| ***7*** | lvPPA | AD | lvPPA | nfvPPA | AD | FTLD-tau CBD | AD |
| ***8*** | lvPPA | AD | lvPPA | nfvPPA | AD | FTLD-tau CBD | AD |
| ***9*** | lvPPA | AD | lvPPA | nfvPPA | AD | FTLD-tau CBD | AD |
| ***10*** | lvPPA | AD | lvPPA | svPPA | AD | FTLD-TDP-C | AD |
| ***11*** | lvPPA | AD | lvPPA | svPPA | AD | FTLD-TDP-C | AD |
| ***12*** | lvPPA | AD | lvPPA | nfvPPA | AD | FTLD-tau CBD | AD |
| ***13*** | lvPPA | AD | lvPPA | nfvPPA | AD | FTLD-tau CBD | AD |
| ***14*** | lvPPA | AD | lvPPA | nfvPPA | AD | FTLD-tau CBD | AD |
| ***15*** | lvPPA | AD | lvPPA | nfvPPA | AD | FTLD-tau CBD | AD |
| ***16*** | lvPPA | AD | lvPPA | svPPA | AD | FTLD-TDP-C | FTLD-TDP |
| ***17*** | lvPPA | AD | lvPPA | nfvPPA | AD | FTLD-tau PSP | AD |
| ***18*** | nfvPPA | FTLD-tau CBD | nfvPPA | lvPPA | FTLD-tau CBD | FTLD-tau PSP | FTLD not specified |
| ***19*** | nfvPPA | FTLD-tau CBD | nfvPPA | lvPPA | FTLD-tau PSP | FTLD-tau CBD | Not specified |
| ***20*** | nfvPPA | FTLD-tau CBD | nfvPPA | lvPPA | FTLD-tau CBD | FTLD-tau PSP | FTLD not specified |
| ***21*** | nfvPPA | FTLD-tau CBD | nfvPPA | lvPPA | FTLD-tau CBD | FTLD-tau PSP | FTLD not specified |
| ***22*** | nfvPPA | FTLD-tau CBD | nfvPPA | lvPPA | FTLD-tau CBD | FTLD-tau PSP | FTLD-tau CBD |
| ***23*** | nfvPPA | FTLD-tau CBD | nfvPPA | lvPPA | FTLD-tau CBD | FTLD-tau PSP | FTLD-tau PSP |
| ***24*** | nfvPPA | FTLD-tau CBD | nfvPPA | lvPPA | FTLD-tau CBD | FTLD-tau PSP | FTLD-tau CBD |
| ***25*** | nfvPPA | FTLD-tau CBD | nfvPPA | lvPPA | FTLD-tau PSP | FTLD-tau CBD | FTLD not specified |
| ***26*** | nfvPPA | FTLD-tau CBD | nfvPPA | lvPPA | FTLD-tau CBD | AD | FTLD-tau PSP |
| ***27*** | nfvPPA | FTLD-tau CBD | nfvPPA | lvPPA | FTLD-tau CBD | FTLD-tau PSP | FTLD-tau CBD |
| ***28*** | nfvPPA | FTLD-tau PSP | nfvPPA | lvPPA | FTLD-tau CBD | FTLD-tau PSP | Not specified |
| ***29*** | nfvPPA | FTLD-tau PSP | nfvPPA | svPPA | FTLD-tau PSP | FTLD-TDP-C | Not specified |
| ***30*** | nfvPPA | FTLD-tau PSP | nfvPPA | lvPPA | FTLD-tau CBD | FTLD-tau PSP | FTLD-tau CBD |
| ***31*** | nfvPPA | FTLD-tau PSP | nfvPPA | lvPPA | FTLD-tau PSP | FTLD-tau CBD | FTLD-tau PSP |
| ***32*** | nfvPPA | FTLD-tau PSP | nfvPPA | lvPPA | FTLD-tau PSP | FTLD-tau CBD | FTLD-tau PSP |
| ***33*** | nfvPPA | FTLD-tau PSP | nfvPPA | lvPPA | FTLD-tau PSP | FTLD-tau CBD | FTLD-tau CBD |
| ***34*** | nfvPPA | FTLD-tau PSP | nfvPPA | lvPPA | FTLD-tau PSP | FTLD-tau CBD | FTLD-tau PSP |
| ***35*** | nfvPPA | FTLD-tau PSP | nfvPPA | lvPPA | FTLD-tau CBD | FTLD-tau PSP | FTLD-tau CBD |
| ***36*** | nfvPPA | FTLD-tau PSP | nfvPPA | lvPPA | FTLD-tau PSP | FTLD-tau CBD | FTLD-TDP |
| ***37*** | svPPA | FTLD-TDP-C | svPPA | lvPPA | FTLD-TDP-C | AD | Not specified |

| | | | | | | | |
|---|---|---|---|---|---|---|---|
| ***38*** | svPPA | FTLD-TDP-C | svPPA | lvPPA | FTLD-TDP-C | AD | Not specified |
| ***39*** | svPPA | FTLD-TDP-C | svPPA | lvPPA | FTLD-TDP-C | AD | Not specified |
| ***40*** | svPPA | FTLD-TDP-C | svPPA | lvPPA | FTLD-TDP-C | AD | Not specified |
| ***41*** | svPPA | FTLD-TDP-C | svPPA | lvPPA | FTLD-TDP-C | AD | Not specified |
| ***42*** | svPPA | FTLD-TDP-C | svPPA | nfvPPA | FTLD-TDP-C | FTLD-tau CBD | FTLD-tau CBD |
| ***43*** | svPPA | FTLD-TDP-C | svPPA | lvPPA | FTLD-TDP-C | AD | Not specified |
| ***44*** | svPPA | FTLD-TDP-C | svPPA | lvPPA | FTLD-TDP-C | AD | FTLD not specified |
| ***45*** | svPPA | FTLD-TDP-C | svPPA | lvPPA | FTLD-TDP-C | AD | Not specified |
| ***46*** | svPPA | FTLD-TDP-C | svPPA | lvPPA | FTLD-TDP-C | AD | FTLD not specified |
| ***47*** | svPPA | FTLD-TDP-C | svPPA | lvPPA | FTLD-TDP-C | AD | FTLD not specified |
| ***48*** | svPPA | FTLD-TDP-C | svPPA | lvPPA | FTLD-TDP-C | AD | Not specified |
| ***49*** | svPPA | FTLD-TDP-C | svPPA | lvPPA | FTLD-TDP-C | AD | FTLD not specified |
| ***50*** | svPPA | FTLD-TDP-C | svPPA | lvPPA | FTLD-TDP-C | AD | FTLD-TDP |
| ***51*** | svPPA | FTLD-TDP-C | svPPA | lvPPA | FTLD-TDP-C | AD | FTLD-TDP |
| ***52*** | svPPA | FTLD-TDP-C | svPPA | lvPPA | FTLD-TDP-C | AD | FTLD not specified |
| ***53*** | svPPA | FTLD-TDP-C | svPPA | lvPPA | FTLD-TDP-C | AD | FTLD-TDP |
| ***54*** | svPPA | FTLD-TDP-C | svPPA | lvPPA | FTLD-TDP-C | AD | FTLD-TDP-C |

**Supplementary Table 2**. Top 20 Differentiating Symptom Clusters by Neuropathological Diagnosis

| NPDx comparison | NPDx 1 | NPDx 2 | rank | cluster_name | % NPDx 1 | % NPDx 2 | % difference | favors NPDx |
|---|---|---|---|---|---|---|---|---|
| **AD_vs_FTLD_TAU-CBD** | AD | FTLD_TAU-CBD | 1 | Progressive Memory Impairment | 10.38 | 3.64 | 6.73 | AD |
| **AD_vs_FTLD_TAU-CBD** | AD | FTLD_TAU-CBD | 2 | Disorientation and Navigational Difficulties | 4.39 | 1.93 | 2.46 | AD |
| **AD_vs_FTLD_TAU-CBD** | AD | FTLD_TAU-CBD | 3 | Neuropsychological Testing: Executive, Memory, and Visuospatial Deficits | 1.46 | 0.15 | 1.31 | AD |
| **AD_vs_FTLD_TAU-CBD** | AD | FTLD_TAU-CBD | 4 | Neuroimaging findings: brain atrophy, white matter changes, and hypometabolism | 1.52 | 0.52 | 1 | AD |
| **AD_vs_FTLD_TAU-CBD** | AD | FTLD_TAU-CBD | 5 | Word-finding difficulties (anomia) | 1.09 | 0.15 | 0.94 | AD |
| **AD_vs_FTLD_TAU-CBD** | AD | FTLD_TAU-CBD | 6 | Distal vibratory sensory loss | 2.12 | 1.26 | 0.85 | AD |
| **AD_vs_FTLD_TAU-CBD** | AD | FTLD_TAU-CBD | 7 | Hallucinations and Illusions (Visual, Auditory, Extracampine) | 1.18 | 0.37 | 0.81 | AD |
| **AD_vs_FTLD_TAU-CBD** | AD | FTLD_TAU-CBD | 8 | Myoclonic Jerks and Involuntary Upper Extremity Movements | 1.09 | 0.3 | 0.8 | AD |
| **AD_vs_FTLD_TAU-CBD** | AD | FTLD_TAU-CBD | 9 | Axial Rigidity | 0.3 | 1.04 | -0.75 | FTLD_TAU-CBD |
| **AD_vs_FTLD_TAU-CBD** | AD | FTLD_TAU-CBD | 10 | Impulsivity and Impulsive Behaviors | 0.18 | 0.97 | -0.78 | FTLD_TAU-CBD |
| **AD_vs_FTLD_TAU-CBD** | AD | FTLD_TAU-CBD | 11 | Social withdrawal and reduced emotional engagement | 0.52 | 1.34 | -0.81 | FTLD_TAU-CBD |
| **AD_vs_FTLD_TAU-CBD** | AD | FTLD_TAU-CBD | 12 | Personality and Behavioral Changes | 0.02 | 0.89 | -0.87 | FTLD_TAU-CBD |
| **AD_vs_FTLD_TAU-CBD** | AD | FTLD_TAU-CBD | 13 | Oral-Buccal (Orofacial) Apraxia | 0.07 | 0.97 | -0.9 | FTLD_TAU-CBD |
| **AD_vs_FTLD_TAU-CBD** | AD | FTLD_TAU-CBD | 14 | Decreased Speech Output | 0.27 | 1.19 | -0.92 | FTLD_TAU-CBD |
| **AD_vs_FTLD_TAU-CBD** | AD | FTLD_TAU-CBD | 15 | Perseveration and Echolalia | 0.18 | 1.12 | -0.93 | FTLD_TAU-CBD |
| **AD_vs_FTLD_TAU-CBD** | AD | FTLD_TAU-CBD | 16 | Cogwheel rigidity and increased muscle tone | 1.3 | 2.23 | -0.93 | FTLD_TAU-CBD |
| **AD_vs_FTLD_TAU-CBD** | AD | FTLD_TAU-CBD | 17 | Bradykinesia and Parkinsonian Motor Symptoms | 0.86 | 1.93 | -1.07 | FTLD_TAU-CBD |
| **AD_vs_FTLD_TAU-CBD** | AD | FTLD_TAU-CBD | 18 | Limb Apraxia | 2.32 | 3.49 | -1.17 | FTLD_TAU-CBD |
| **AD_vs_FTLD_TAU-CBD** | AD | FTLD_TAU-CBD | 19 | Repetitive and Compulsive Behaviors with Cognitive Rigidity | 0.84 | 2.23 | -1.39 | FTLD_TAU-CBD |
| **AD_vs_FTLD_TAU-CBD** | AD | FTLD_TAU-CBD | 20 | Progressive unilateral arm/hand weakness and abnormal movements | 0.23 | 2.23 | -2 | FTLD_TAU-CBD |
| **AD_vs_FTLD_TAU-PSP** | AD | FTLD_TAU-PSP | 1 | Progressive Memory Impairment | 10.25 | 2.65 | 7.6 | AD |
| **AD_vs_FTLD_TAU-PSP** | AD | FTLD_TAU-PSP | 2 | Disorientation and Navigational Difficulties | 4.34 | 0.71 | 3.63 | AD |
| **AD_vs_FTLD_TAU-PSP** | AD | FTLD_TAU-PSP | 3 | Increased Irritability, Anger, and Behavioral Changes | 2.67 | 0.85 | 1.82 | AD |
| **AD_vs_FTLD_TAU-PSP** | AD | FTLD_TAU-PSP | 4 | Visual Field Deficits and Hemispatial Neglect | 1.21 | 0.09 | 1.12 | AD |
| **AD_vs_FTLD_TAU-PSP** | AD | FTLD_TAU-PSP | 5 | Frequent misplacing or losing objects | 1.21 | 0.14 | 1.07 | AD |
| **AD_vs_FTLD_TAU-PSP** | AD | FTLD_TAU-PSP | 6 | Hallucinations and Illusions (Visual, Auditory, Extracampine) | 1.17 | 0.14 | 1.03 | AD |
| **AD_vs_FTLD_TAU-PSP** | AD | FTLD_TAU-PSP | 7 | Neuroimaging findings: brain atrophy, white matter changes, and hypometabolism | 1.51 | 0.57 | 0.94 | AD |
| **AD_vs_FTLD_TAU-PSP** | AD | FTLD_TAU-PSP | 8 | Increased Anxiety and Panic Symptoms | 1.44 | 0.52 | 0.92 | AD |

| | | | | | | | | |
|---|---|---|---|---|---|---|---|---|
| **AD_vs_FTLD_TAU-PSP** | AD | FTLD_TAU-PSP | 9 | Progressive Cognitive Decline and Language Impairment | 1.46 | 0.57 | 0.89 | AD |
| **AD_vs_FTLD_TAU-PSP** | AD | FTLD_TAU-PSP | 10 | Mild Hypomimia (Reduced Facial Expression) | 0.43 | 1.28 | -0.85 | FTLD_TAU-PSP |
| **AD_vs_FTLD_TAU-PSP** | AD | FTLD_TAU-PSP | 11 | Upper Extremity Dystonia | 0.29 | 1.23 | -0.94 | FTLD_TAU-PSP |
| **AD_vs_FTLD_TAU-PSP** | AD | FTLD_TAU-PSP | 12 | Square wave jerks and abnormal saccadic eye movements | 0.04 | 1.04 | -1 | FTLD_TAU-PSP |
| **AD_vs_FTLD_TAU-PSP** | AD | FTLD_TAU-PSP | 13 | Emotional Lability and Pseudobulbar Affect | 0.92 | 1.94 | -1.02 | FTLD_TAU-PSP |
| **AD_vs_FTLD_TAU-PSP** | AD | FTLD_TAU-PSP | 14 | Masked Facies and Parkinsonian Facial Features | 0.29 | 1.65 | -1.36 | FTLD_TAU-PSP |
| **AD_vs_FTLD_TAU-PSP** | AD | FTLD_TAU-PSP | 15 | Impaired Extraocular Movements (Especially Vertical Gaze Palsy) | 0.27 | 1.84 | -1.57 | FTLD_TAU-PSP |
| **AD_vs_FTLD_TAU-PSP** | AD | FTLD_TAU-PSP | 16 | Slow and Impaired Saccadic Eye Movements | 0.67 | 2.41 | -1.74 | FTLD_TAU-PSP |
| **AD_vs_FTLD_TAU-PSP** | AD | FTLD_TAU-PSP | 17 | Chronic Visual Disturbances (e.g., diplopia, blurred vision, depth perception problems) | 0.43 | 2.17 | -1.75 | FTLD_TAU-PSP |
| **AD_vs_FTLD_TAU-PSP** | AD | FTLD_TAU-PSP | 18 | Axial Rigidity | 0.29 | 2.41 | -2.12 | FTLD_TAU-PSP |
| **AD_vs_FTLD_TAU-PSP** | AD | FTLD_TAU-PSP | 19 | Bradykinesia and Parkinsonian Motor Symptoms | 0.85 | 3.12 | -2.27 | FTLD_TAU-PSP |
| **AD_vs_FTLD_TAU-PSP** | AD | FTLD_TAU-PSP | 20 | Dysphagia and Swallowing Difficulties | 0.56 | 4.4 | -3.84 | FTLD_TAU-PSP |
| **AD_vs_FTLD_TAU-TDP-C** | AD | FTLD_TAU-TDP-C | 1 | Progressive Memory Impairment | 10.41 | 6.02 | 4.39 | AD |
| **AD_vs_FTLD_TAU-TDP-C** | AD | FTLD_TAU-TDP-C | 2 | Disorientation and Navigational Difficulties | 4.41 | 1.61 | 2.79 | AD |
| **AD_vs_FTLD_TAU-TDP-C** | AD | FTLD_TAU-TDP-C | 3 | Limb Apraxia | 2.33 | 0.21 | 2.11 | AD |
| **AD_vs_FTLD_TAU-TDP-C** | AD | FTLD_TAU-TDP-C | 4 | Impaired Rapid Repetitive Movements (Finger/Toe Tapping) | 2.53 | 0.75 | 1.78 | AD |
| **AD_vs_FTLD_TAU-TDP-C** | AD | FTLD_TAU-TDP-C | 5 | Upper Extremity Tremor (Rest, Postural, and Action) | 3.26 | 1.5 | 1.76 | AD |
| **AD_vs_FTLD_TAU-TDP-C** | AD | FTLD_TAU-TDP-C | 6 | Visual Field Deficits and Hemispatial Neglect | 1.23 | 0 | 1.23 | AD |
| **AD_vs_FTLD_TAU-TDP-C** | AD | FTLD_TAU-TDP-C | 7 | Increased Irritability, Anger, and Behavioral Changes | 2.72 | 1.61 | 1.11 | AD |
| **AD_vs_FTLD_TAU-TDP-C** | AD | FTLD_TAU-TDP-C | 8 | Myoclonic Jerks and Involuntary Upper Extremity Movements | 1.1 | 0 | 1.1 | AD |
| **AD_vs_FTLD_TAU-TDP-C** | AD | FTLD_TAU-TDP-C | 9 | Progressive Loss of Empathy and Social Awareness | 0.07 | 1.18 | -1.11 | FTLD_TAU-TDP-C |
| **AD_vs_FTLD_TAU-TDP-C** | AD | FTLD_TAU-TDP-C | 10 | Decreased Speech Output | 0.27 | 1.4 | -1.12 | FTLD_TAU-TDP-C |
| **AD_vs_FTLD_TAU-TDP-C** | AD | FTLD_TAU-TDP-C | 11 | Surface Dyslexia | 0.05 | 1.18 | -1.14 | FTLD_TAU-TDP-C |
| **AD_vs_FTLD_TAU-TDP-C** | AD | FTLD_TAU-TDP-C | 12 | Personality and Behavioral Changes | 0.23 | 1.4 | -1.17 | FTLD_TAU-TDP-C |
| **AD_vs_FTLD_TAU-TDP-C** | AD | FTLD_TAU-TDP-C | 13 | Emotional blunting and apathy | 0.09 | 1.29 | -1.2 | FTLD_TAU-TDP-C |
| **AD_vs_FTLD_TAU-TDP-C** | AD | FTLD_TAU-TDP-C | 14 | Progressive word-finding and language expression difficulties | 0.84 | 2.15 | -1.3 | FTLD_TAU-TDP-C |
| **AD_vs_FTLD_TAU-TDP-C** | AD | FTLD_TAU-TDP-C | 15 | Apathy and Decreased Motivation | 1.14 | 2.47 | -1.33 | FTLD_TAU-TDP-C |
| **AD_vs_FTLD_TAU-TDP-C** | AD | FTLD_TAU-TDP-C | 16 | Increased Sweet Cravings and Overeating Leading to Weight Gain | 0.39 | 2.04 | -1.65 | FTLD_TAU-TDP-C |
| **AD_vs_FTLD_TAU-TDP-C** | AD | FTLD_TAU-TDP-C | 17 | Flat or Restricted Affect | 0.59 | 2.47 | -1.88 | FTLD_TAU-TDP-C |
| **AD_vs_FTLD_TAU-TDP-C** | AD | FTLD_TAU-TDP-C | 18 | Behavioral disinhibition and socially inappropriate behavior | 1.23 | 4.51 | -3.28 | FTLD_TAU-TDP-C |
| **AD_vs_FTLD_TAU-TDP-C** | AD | FTLD_TAU-TDP-C | 19 | Difficulty Recognizing Faces (Prosopagnosia) | 0.5 | 3.97 | -3.47 | FTLD_TAU-TDP-C |
| **AD_vs_FTLD_TAU-TDP-C** | AD | FTLD_TAU-TDP-C | 20 | Repetitive and Compulsive Behaviors with Cognitive Rigidity | 0.84 | 8.59 | -7.75 | FTLD_TAU-TDP-C |
| **FTLD_TAU-CBD_vs_FTLD_TAU-TDP-C** | FTLD_TAU-CBD | FTLD_TAU-TDP-C | 1 | Limb Apraxia | 4.15 | 0.25 | 3.9 | FTLD_TAU-CBD |
| **FTLD_TAU-CBD_vs_FTLD_TAU-TDP-C** | FTLD_TAU-CBD | FTLD_TAU-TDP-C | 2 | Upper Extremity Tremor (Rest, Postural, and Action) | 4.41 | 1.72 | 2.69 | FTLD_TAU-CBD |
| **FTLD_TAU-CBD_vs_FTLD_TAU-TDP-C** | FTLD_TAU-CBD | FTLD_TAU-TDP-C | 3 | Progressive unilateral arm/hand weakness and abnormal movements | 2.65 | 0 | 2.65 | FTLD_TAU-CBD |
| **FTLD_TAU-CBD_vs_FTLD_TAU-TDP-C** | FTLD_TAU-CBD | FTLD_TAU-TDP-C | 4 | Cogwheel rigidity and increased muscle tone | 2.65 | 0.62 | 2.03 | FTLD_TAU-CBD |
| **FTLD_TAU-CBD_vs_FTLD_TAU-TDP-C** | FTLD_TAU-CBD | FTLD_TAU-TDP-C | 5 | Impaired Rapid Repetitive Movements (Finger/Toe Tapping) | 2.65 | 0.86 | 1.79 | FTLD_TAU-CBD |
| **FTLD_TAU-CBD_vs_FTLD_TAU-TDP-C** | FTLD_TAU-CBD | FTLD_TAU-TDP-C | 6 | Bradykinesia and Parkinsonian Motor Symptoms | 2.29 | 0.62 | 1.68 | FTLD_TAU-CBD |
| **FTLD_TAU-CBD_vs_FTLD_TAU-TDP-C** | FTLD_TAU-CBD | FTLD_TAU-TDP-C | 7 | Perseveration and Echolalia | 1.32 | 0 | 1.32 | FTLD_TAU-CBD |
| **FTLD_TAU-CBD_vs_FTLD_TAU-TDP-C** | FTLD_TAU-CBD | FTLD_TAU-TDP-C | 8 | Parkinsonian gait | 2.91 | 1.6 | 1.31 | FTLD_TAU-CBD |
| **FTLD_TAU-CBD_vs_FTLD_TAU-TDP-C** | FTLD_TAU-CBD | FTLD_TAU-TDP-C | 9 | Orthostatic Hypotension and Lightheadedness | 1.5 | 0.25 | 1.25 | FTLD_TAU-CBD |
| **FTLD_TAU-CBD_vs_FTLD_TAU-TDP-C** | FTLD_TAU-CBD | FTLD_TAU-TDP-C | 10 | Axial Rigidity | 1.24 | 0 | 1.24 | FTLD_TAU-CBD |
| **FTLD_TAU-CBD_vs_FTLD_TAU-TDP-C** | FTLD_TAU-CBD | FTLD_TAU-TDP-C | 11 | Peripheral Neuropathy and Limb Paresthesia | 1.24 | 0 | 1.24 | FTLD_TAU-CBD |
| **FTLD_TAU-CBD_vs_FTLD_TAU-TDP-C** | FTLD_TAU-CBD | FTLD_TAU-TDP-C | 12 | Oral-Buccal (Orofacial) Apraxia | 1.15 | 0 | 1.15 | FTLD_TAU-CBD |
| **FTLD_TAU-CBD_vs_FTLD_TAU-TDP-C** | FTLD_TAU-CBD | FTLD_TAU-TDP-C | 13 | Surface Dyslexia | 0.09 | 1.35 | -1.26 | FTLD_TAU-TDP-C |
| **FTLD_TAU-CBD_vs_FTLD_TAU-TDP-C** | FTLD_TAU-CBD | FTLD_TAU-TDP-C | 14 | Increased Sweet Cravings and Overeating Leading to Weight Gain | 0.71 | 2.34 | -1.63 | FTLD_TAU-TDP-C |
| **FTLD_TAU-CBD_vs_FTLD_TAU-TDP-C** | FTLD_TAU-CBD | FTLD_TAU-TDP-C | 15 | Progressive word-finding and language expression difficulties | 0.44 | 2.46 | -2.02 | FTLD_TAU-TDP-C |
| **FTLD_TAU-CBD_vs_FTLD_TAU-TDP-C** | FTLD_TAU-CBD | FTLD_TAU-TDP-C | 16 | Flat or Restricted Affect | 0.71 | 2.83 | -2.12 | FTLD_TAU-TDP-C |
| **FTLD_TAU-CBD_vs_FTLD_TAU-TDP-C** | FTLD_TAU-CBD | FTLD_TAU-TDP-C | 17 | Progressive Memory Impairment | 4.32 | 6.89 | -2.56 | FTLD_TAU-TDP-C |
| **FTLD_TAU-CBD_vs_FTLD_TAU-TDP-C** | FTLD_TAU-CBD | FTLD_TAU-TDP-C | 18 | Behavioral disinhibition and socially inappropriate behavior | 1.94 | 5.17 | -3.22 | FTLD_TAU-TDP-C |
| **FTLD_TAU-CBD_vs_FTLD_TAU-TDP-C** | FTLD_TAU-CBD | FTLD_TAU-TDP-C | 19 | Difficulty Recognizing Faces (Prosopagnosia) | 0.18 | 4.55 | -4.37 | FTLD_TAU-TDP-C |
| **FTLD_TAU-CBD_vs_FTLD_TAU-TDP-C** | FTLD_TAU-CBD | FTLD_TAU-TDP-C | 20 | Repetitive and Compulsive Behaviors with Cognitive Rigidity | 2.65 | 9.84 | -7.19 | FTLD_TAU-TDP-C |
| **FTLD_TAU-PSP_vs_FTLD_TAU-CBD** | FTLD_TAU-PSP | FTLD_TAU-CBD | 1 | Dysphagia and Swallowing Difficulties | 4.58 | 0.91 | 3.67 | FTLD_TAU-PSP |
| **FTLD_TAU-PSP_vs_FTLD_TAU-CBD** | FTLD_TAU-PSP | FTLD_TAU-CBD | 2 | Chronic Visual Disturbances (e.g., diplopia, blurred vision, depth perception problems) | 2.27 | 0.15 | 2.11 | FTLD_TAU-PSP |
| **FTLD_TAU-PSP_vs_FTLD_TAU-CBD** | FTLD_TAU-PSP | FTLD_TAU-CBD | 3 | Slow and Impaired Saccadic Eye Movements | 2.51 | 0.91 | 1.6 | FTLD_TAU-PSP |
| **FTLD_TAU-PSP_vs_FTLD_TAU-CBD** | FTLD_TAU-PSP | FTLD_TAU-CBD | 4 | Axial Rigidity | 2.51 | 1.07 | 1.45 | FTLD_TAU-PSP |
| **FTLD_TAU-PSP_vs_FTLD_TAU-CBD** | FTLD_TAU-PSP | FTLD_TAU-CBD | 5 | Masked Facies and Parkinsonian Facial Features | 1.72 | 0.3 | 1.42 | FTLD_TAU-PSP |
| **FTLD_TAU-PSP_vs_FTLD_TAU-CBD** | FTLD_TAU-PSP | FTLD_TAU-CBD | 6 | Impaired Extraocular Movements (Especially Vertical Gaze Palsy) | 1.92 | 0.61 | 1.31 | FTLD_TAU-PSP |
| **FTLD_TAU-PSP_vs_FTLD_TAU-CBD** | FTLD_TAU-PSP | FTLD_TAU-CBD | 7 | Bradykinesia and Parkinsonian Motor Symptoms | 3.25 | 1.98 | 1.27 | FTLD_TAU-PSP |

| | | | | | | | | |
|---|---|---|---|---|---|---|---|---|
| **FTLD_TAU-PSP_vs_FTLD_TAU-CBD** | FTLD_TAU-PSP | FTLD_TAU-CBD | 8 | Impaired Rapid Repetitive Movements (Finger/Toe Tapping) | 3.35 | 2.28 | 1.07 | FTLD_TAU-PSP |
| **FTLD_TAU-PSP_vs_FTLD_TAU-CBD** | FTLD_TAU-PSP | FTLD_TAU-CBD | 9 | Emotional Lability and Pseudobulbar Affect | 2.02 | 1.07 | 0.95 | FTLD_TAU-PSP |
| **FTLD_TAU-PSP_vs_FTLD_TAU-CBD** | FTLD_TAU-PSP | FTLD_TAU-CBD | 10 | Cerebellar Coordination Deficits (e.g., Dysmetria, Dysdiadochokinesis, Ataxia) | 0.15 | 1.07 | -0.92 | FTLD_TAU-CBD |
| **FTLD_TAU-PSP_vs_FTLD_TAU-CBD** | FTLD_TAU-PSP | FTLD_TAU-CBD | 11 | Cogwheel rigidity and increased muscle tone | 1.33 | 2.28 | -0.95 | FTLD_TAU-CBD |
| **FTLD_TAU-PSP_vs_FTLD_TAU-CBD** | FTLD_TAU-PSP | FTLD_TAU-CBD | 12 | Progressive Memory Impairment | 2.76 | 3.73 | -0.97 | FTLD_TAU-CBD |
| **FTLD_TAU-PSP_vs_FTLD_TAU-CBD** | FTLD_TAU-PSP | FTLD_TAU-CBD | 13 | Speech Disturbances (Effortful, Halting, Paraphasic, Word-Finding Difficulties) | 1.38 | 2.36 | -0.98 | FTLD_TAU-CBD |
| **FTLD_TAU-PSP_vs_FTLD_TAU-CBD** | FTLD_TAU-PSP | FTLD_TAU-CBD | 14 | Perseveration and Echolalia | 0.15 | 1.14 | -0.99 | FTLD_TAU-CBD |
| **FTLD_TAU-PSP_vs_FTLD_TAU-CBD** | FTLD_TAU-PSP | FTLD_TAU-CBD | 15 | Repetitive and Compulsive Behaviors with Cognitive Rigidity | 1.23 | 2.28 | -1.05 | FTLD_TAU-CBD |
| **FTLD_TAU-PSP_vs_FTLD_TAU-CBD** | FTLD_TAU-PSP | FTLD_TAU-CBD | 16 | Behavioral disinhibition and socially inappropriate behavior | 0.59 | 1.67 | -1.08 | FTLD_TAU-CBD |
| **FTLD_TAU-PSP_vs_FTLD_TAU-CBD** | FTLD_TAU-PSP | FTLD_TAU-CBD | 17 | Disorientation and Navigational Difficulties | 0.74 | 1.98 | -1.24 | FTLD_TAU-CBD |
| **FTLD_TAU-PSP_vs_FTLD_TAU-CBD** | FTLD_TAU-PSP | FTLD_TAU-CBD | 18 | Progressive unilateral arm/hand weakness and abnormal movements | 0.84 | 2.28 | -1.45 | FTLD_TAU-CBD |
| **FTLD_TAU-PSP_vs_FTLD_TAU-CBD** | FTLD_TAU-PSP | FTLD_TAU-CBD | 19 | Increased Irritability, Anger, and Behavioral Changes | 0.89 | 2.51 | -1.62 | FTLD_TAU-CBD |
| **FTLD_TAU-PSP_vs_FTLD_TAU-CBD** | FTLD_TAU-PSP | FTLD_TAU-CBD | 20 | Limb Apraxia | 1.72 | 3.58 | -1.85 | FTLD_TAU-CBD |
| **FTLD_TAU-PSP_vs_FTLD_TAU-TDP-C** | FTLD_TAU-PSP | FTLD_TAU-TDP-C | 1 | Dysphagia and Swallowing Difficulties | 4.76 | 0.94 | 3.82 | FTLD_TAU-PSP |
| **FTLD_TAU-PSP_vs_FTLD_TAU-TDP-C** | FTLD_TAU-PSP | FTLD_TAU-TDP-C | 2 | Bradykinesia and Parkinsonian Motor Symptoms | 3.38 | 0.59 | 2.79 | FTLD_TAU-PSP |
| **FTLD_TAU-PSP_vs_FTLD_TAU-TDP-C** | FTLD_TAU-PSP | FTLD_TAU-TDP-C | 3 | Impaired Rapid Repetitive Movements (Finger/Toe Tapping) | 3.48 | 0.82 | 2.65 | FTLD_TAU-PSP |
| **FTLD_TAU-PSP_vs_FTLD_TAU-TDP-C** | FTLD_TAU-PSP | FTLD_TAU-TDP-C | 4 | Slow and Impaired Saccadic Eye Movements | 2.61 | 0 | 2.61 | FTLD_TAU-PSP |
| **FTLD_TAU-PSP_vs_FTLD_TAU-TDP-C** | FTLD_TAU-PSP | FTLD_TAU-TDP-C | 5 | Axial Rigidity | 2.61 | 0 | 2.61 | FTLD_TAU-PSP |
| **FTLD_TAU-PSP_vs_FTLD_TAU-TDP-C** | FTLD_TAU-PSP | FTLD_TAU-TDP-C | 6 | Upper Extremity Tremor (Rest, Postural, and Action) | 3.99 | 1.65 | 2.34 | FTLD_TAU-PSP |
| **FTLD_TAU-PSP_vs_FTLD_TAU-TDP-C** | FTLD_TAU-PSP | FTLD_TAU-TDP-C | 7 | Chronic Visual Disturbances (e.g., diplopia, blurred vision, depth perception problems) | 2.35 | 0.24 | 2.12 | FTLD_TAU-PSP |
| **FTLD_TAU-PSP_vs_FTLD_TAU-TDP-C** | FTLD_TAU-PSP | FTLD_TAU-TDP-C | 8 | Impaired Extraocular Movements (Especially Vertical Gaze Palsy) | 1.99 | 0.24 | 1.76 | FTLD_TAU-PSP |
| **FTLD_TAU-PSP_vs_FTLD_TAU-TDP-C** | FTLD_TAU-PSP | FTLD_TAU-TDP-C | 9 | Limb Apraxia | 1.79 | 0.24 | 1.55 | FTLD_TAU-PSP |
| **FTLD_TAU-PSP_vs_FTLD_TAU-TDP-C** | FTLD_TAU-PSP | FTLD_TAU-TDP-C | 10 | Masked Facies and Parkinsonian Facial Features | 1.79 | 0.24 | 1.55 | FTLD_TAU-PSP |
| **FTLD_TAU-PSP_vs_FTLD_TAU-TDP-C** | FTLD_TAU-PSP | FTLD_TAU-TDP-C | 11 | Surface Dyslexia | 0 | 1.29 | -1.29 | FTLD_TAU-TDP-C |
| **FTLD_TAU-PSP_vs_FTLD_TAU-TDP-C** | FTLD_TAU-PSP | FTLD_TAU-TDP-C | 12 | Paraphasic and Phonemic Speech Errors | 0.61 | 2.24 | -1.62 | FTLD_TAU-TDP-C |
| **FTLD_TAU-PSP_vs_FTLD_TAU-TDP-C** | FTLD_TAU-PSP | FTLD_TAU-TDP-C | 13 | Language and Reading Comprehension Difficulties | 0.26 | 1.88 | -1.63 | FTLD_TAU-TDP-C |
| **FTLD_TAU-PSP_vs_FTLD_TAU-TDP-C** | FTLD_TAU-PSP | FTLD_TAU-TDP-C | 14 | Progressive word-finding and language expression difficulties | 0.72 | 2.35 | -1.64 | FTLD_TAU-TDP-C |
| **FTLD_TAU-PSP_vs_FTLD_TAU-TDP-C** | FTLD_TAU-PSP | FTLD_TAU-TDP-C | 15 | Increased Sweet Cravings and Overeating Leading to Weight Gain | 0.31 | 2.24 | -1.93 | FTLD_TAU-TDP-C |
| **FTLD_TAU-PSP_vs_FTLD_TAU-TDP-C** | FTLD_TAU-PSP | FTLD_TAU-TDP-C | 16 | Flat or Restricted Affect | 0.66 | 2.71 | -2.04 | FTLD_TAU-TDP-C |
| **FTLD_TAU-PSP_vs_FTLD_TAU-TDP-C** | FTLD_TAU-PSP | FTLD_TAU-TDP-C | 17 | Progressive Memory Impairment | 2.86 | 6.59 | -3.72 | FTLD_TAU-TDP-C |
| **FTLD_TAU-PSP_vs_FTLD_TAU-TDP-C** | FTLD_TAU-PSP | FTLD_TAU-TDP-C | 18 | Difficulty Recognizing Faces (Prosopagnosia) | 0.15 | 4.35 | -4.2 | FTLD_TAU-TDP-C |
| **FTLD_TAU-PSP_vs_FTLD_TAU-TDP-C** | FTLD_TAU-PSP | FTLD_TAU-TDP-C | 19 | Behavioral disinhibition and socially inappropriate behavior | 0.61 | 4.94 | -4.33 | FTLD_TAU-TDP-C |
| **FTLD_TAU-PSP_vs_FTLD_TAU-TDP-C** | FTLD_TAU-PSP | FTLD_TAU-TDP-C | 20 | Repetitive and Compulsive Behaviors with Cognitive Rigidity | 1.28 | 9.41 | -8.13 | FTLD_TAU-TDP-C |

**Brain image processing and W-maps creation**

Structural T1-weigthed images were preprocessed using SPM12. The images were visually inspected for artifacts, and underwent bias-correction, segmentation into tissue compartments, and spatial normalization using a single generative model with the standard SPM12 parameters. The default tissue probability maps for grey matter, white matter, cerebrospinal fluid, and all other voxels from SPM12 (TPM.nii) were used[1]. To optimize intersubject registration, each participant's image was warped to a template derived from 300 confirmed neurologically healthy older adults (ages 44-86, M±SD: 67.2±7.3; 113 males, 186 females) scanned with one of three magnet strengths (1.5T, 3T, 4T), using affine and nonlinear transformations with the help of the diffeomorphic anatomical registration through exponentiated lie algebra (DARTEL) method, with standard implementation in SPM12[1]. In all preprocessing steps, default parameters of the SPM12 toolbox were used. Total volume of each tissue compartment was calculated by applying the modulated, warped and segmented masks for gray matter, white matter, and CSF to the corresponding MWS probability map for that individual, and the total intracranial volume (TIV) was derived by summing the three volumes. The spatially normalized, segmented, and modulated gray matter images were smoothed with an 8-mm FWHM isotropic Gaussian kernel.

W-maps were calculated by first performing voxel-wise regressions in SPM12 on the nuisance factors (age, sex, TIV, and magnet field strength) in a set of 534 confirmed neurologically healthy older controls (ages 44-99, M±SD: 68.7±9.1; 220 males, 302 females) from the UCSF MAC Hilblom Cohort to derive beta maps estimating each factor's effect on the gray matter probability images. Then, voxel-wise w-maps for each participant were computed on their smoothed/modulated/warped (SMW) gray matter probability images with the formula: W-score = (actual voxel intensity – expected voxel intensity)/SD. Actual = SMW GM probability for this participant at a given voxel. Expected = the predicted GM probability for that voxel applying the beta weights from the 4 nuisance factors derived from the healthy controls. SD = the standard deviation of the residuals for that voxel among the controls. Thus, the W-score for a voxel represents where that voxel falls on a normal probability distribution of gray matter volume after the 4 nuisance factors are taken into account[2,3]. Negative W-scores represent below-average volume. <-1.50 are below 7th %ile compared to healthy controls and might be considered clinically abnormal. These analyses were performed using the Brainsight system, developed at UCSF by Katherine P. Rankin, Cosmo Mielke, and Paul Sukhanov, and powered by the VLSM script written by Stephen M. Wilson, with funding from the Rainwater Charitable Foundation and the UCSF Chancellor's Fund for Precision Medicine.

**Prompt engineering for the AI agents**

The first step of the AI system multi-agent architecture implemented three specialized LLM agents – modeled after a behavioral neurologist, a neuropsychologist, and a neuroimaging specialist – which processed multimodal input data in parallel, each guided by a distinct system prompt reflecting domain-specific expertise.

*Behavioral Neurologist Agent*

This agent specialized in in clinical syndrome recognition and symptom pattern analysis across neurodegenerative diseases. The agent's system prompt defined its role as a behavioral neurologist with "*extensive expertise in recognizing clinical syndromes based on symptom patterns, progression, and manifestations.*" For the PPA-specific analysis, this agent focused primarily on language and communication features, including characterization of language symptoms, speech characteristics (apraxia of speech, dysarthria), word finding abilities, grammatical processing, comprehension skills, and reading/writing abilities. The agent was instructed to analyze language features, behavioral symptoms, motor features, and clinical progression while explicitly avoiding specific diagnostic labels in favor of detailed characterization.

*Neuropsychologist Agent*

The neuropsychologist agent specialized in cognitive assessment and interpretation of neuropsychological test patterns. Its system prompt established expertise in "*interpreting language and cognitive test patterns, identifying* *domain-specific deficits, and understanding their functional implications.*" The agent performed comprehensive analysis of language profiles including naming and word retrieval, grammar and syntax, comprehension abilities, repetition skills, and speech production. Beyond language, it evaluated memory systems (episodic, semantic, working memory), executive functions (planning, inhibition, cognitive flexibility), attention and processing speed, and visuospatial abilities. The agent was tasked with pattern significance interpretation and cognitive trajectory assessment while focusing solely on describing cognitive profiles without diagnostic labeling.

*Neuroimaging Agent*

The neuroimaging agent specialized in neuroanatomical analysis and structure-function relationships. The system prompt defined its expertise in "*interpreting structural brain changes in language networks, understanding structure-function relationships in language processing, and identifying pathological patterns.*" For language network analysis, the agent examined atrophy patterns in critical language regions including the left inferior frontal gyrus, superior temporal gyrus, middle temporal gyrus, anterior temporal lobe, temporoparietal junction, and insula. The agent assessed hemispheric asymmetry, analyzed structure-function implications of observed atrophy patterns, and evaluated anatomical findings for their relevance to PPA variant differentiation.

**Context engineering for the AI the Board Review Agent**

*Diagnostic Criteria Integration*

The board review agents had access to formal diagnostic criteria for all clinical syndromes through an integrated diagnostic criteria loading system, with documents curated by the clinicians at the UCSF MAC. For PPA variants, the system accessed the established diagnostic criteria defining core features for each variant, curated by the clinicians at the UCSF MAC. The data source covered 15 clinical syndromes: Primary Progressive Aphasia[4], Alzheimer's Disease[5], Amyotrophic Lateral Sclerosis[6], Behavioral Variant Frontotemporal Dementia[7], Corticobasal Degeneration/Syndrome[8], Creutzfeldt-Jakob Disease[9–11], Dementia with Lewy Bodies[12], HIV-Associated Neurocognitive Disorder[13], Ischemic Vascular Dementia[14], Mild Cognitive Impairment[15], Multiple System Atrophy[16], Parkinson's Disease[17], Posterior Cortical Atrophy[18], Progressive Supranuclear Palsy[19] and Traumatic Encephalopathy Syndrome[20]. All diagnostic criteria were dynamically loaded during the board review to ensure adherence to established clinical standards.

*Empirical Differential Diagnosis Database*

We began by assembling the dataset from the complete UCSF MAC Brain Bank cohort, comprising 855 patients diagnosed with a variety of neurodegenerative diseases. We then excluded the 54 patients analyzed in our main results to prevent data leakage. For all included patients, we utilized the entirety of their clinical notes across all available visits. Clinical notes were filtered and selected for information extraction using a structured prompt with OpenAI's gpt-4.1 (version 2025-04-14; temperature=0) via the Microsoft Azure OpenAI service operated through the UCSF Versa API, a secure enterprise generative AI platform approved for P4 identifiable data. The engineered prompt (previously described by our team[21]) directed the model to extract information into a structured JSON schema, enforced with Pydantic, spanning diagnostic codes, clinical findings, structured measurements, medications, and medical history, with special emphasis on clinical findings due to their absence in structured data. Supporting evidence text for each clinical finding was collected and preprocessed through a custom NLP pipeline to standardize text format, including lowercasing, Unicode to ASCII normalization, selective character removal, and whitespace cleanup. The cleaned symptom texts were then embedded as 3072-dimensional vectors using the Azure OpenAI text-embedding-3-large-1 model via Versa, reduced to 100 dimensions by PCA for computational tractability, and visualized in two dimensions using UMAP. Symptom embeddings were clustered using HDBscan with grid search over key parameters, excluding noise points. For each cluster, 15 example supporting evidence snippets were passed to GPT-4o to generate concise, descriptive cluster labels, which were parsed and aggregated into a structured JSON associating each cluster with its label. Each patient's symptoms were then represented as frequency counts of findings by symptom cluster. To construct the empirical differential diagnosis database, patients were grouped by confirmed neuropathological diagnosis (AD, FTLD-TDP-C, FTLD-4R-TAU PSP, FTLD-4R-TAU CBD), and for each pairwise diagnostic comparison, the average frequencies of each symptom cluster across diagnostic groups were computed. Symptom clusters were then ranked according to their absolute percentage differences between groups, producing for each comparison a list of the top 20 clinical symptom clusters that most effectively distinguished the neuropathologies, thereby enabling a quantitative, evidence-based resource for differential diagnosis.

**Technical Implementation**

*Large Language Model Infrastructure and Data Security*

All AI agents utilized OpenAI's o4-mini with high reasoning effort and greddy decoding (temperature set to 0) to ensure the most possible deterministic outputs. Processing occurred entirely within the UCSF Versa platform. The platform also provided comprehensive audit logging to support research reproducibility and regulatory compliance. The framework followed model-agnostic design principles, enabling accommodation of alternative LLMs or large multimodal language models as they become available. *Structured Output Validation:* All AI agent responses underwent structured validation using Pydantic schemas to ensure consistent output formats and completeness. Each agent type had corresponding response schemas defining required fields, data types, and validation rules for diagnostic outputs. *Parallel Processing Architecture:* The system implemented parallel processing for specialist agent assessments using ThreadPoolExecutor with three concurrent workers, enabling simultaneous analysis by behavioral neurologist, neuropsychologist, and neuropathologist agents. This architecture reduced overall processing time while maintaining independence of specialist assessments prior to integration.
